# Prescription opioid-related alterations to amygdalar and thalamic functional networks in chronic knee pain: A retrospective case control resting-state connectivity study

**DOI:** 10.1101/2022.03.01.22271607

**Authors:** Marianne Marta Drabek, William Joseph Cottam, Sarina Jennifer Iwabuchi, Arman Tadjibaev, Ali-Reza Mohammadi-Nejad, Dorothee P Auer

## Abstract

**Objective:** Long-term opioid use is associated with diminished pain relief, hyperalgesia, and addiction which is not well understood. This study aimed to characterise opioid-related brain network alterations in chronic pain, focused on the right amygdala, and left mediodorsal thalamic nuclei that play key roles in affective pain processing, and are particularly rich in mu opioid receptors (MOR).

**Subjects:** Participants on opioid prescriptions with painful knee osteoarthritis and matched non-opioid using control pain participants.

**Methods and design:** Seed-based functional connectivity (FC) maps from resting-state fMRI data were compared between groups.

**Results:** We found right amygdala hyperconnectivity with the posterior default mode network (pDMN) and the dorsomedial prefrontal cortex in opioid users in contrast to anti-correlations in controls. Conversely, opioid users showed predominant hypoconnectivity of the left dorsomedial thalamic seed with the cingulate cortex except for the subgenual part displaying an anti-correlation in opioid users and no association in non-users. Opioid users also showed higher negative affect in exploratory post-hoc tests suggesting a potential contribution of trait anxiety to amygdala-pDMN FC alteration.

**Conclusion:** Opioid use related hyperconnectivity of the right amygdalar network likely reflects maladaptive mechanisms involving negative affect and network plasticity. Hypoconnectivity of the mediodorsal thalamic nuclei with the anterior and mid cingulate on the other hand may reflect impaired resilience in line with previously reported compensatory MOR upregulation. In conclusion, this study provides new insight into possible brain mechanisms underlying adverse effects of prolonged opioids in chronic pain and offer candidate network targets for novel interventions.

## Introduction

Prolonged use of prescription opioids comes with reduced efficacy for pain relief, even paradoxical opioid-induced hyperalgesia, sleep and endocrine impairment [1] and often addiction, further opioid use, and fatalities. Given that most opioid prescriptions are for pain treatment, the opioid crisis can only be tackled with better understanding of opioid and pain interactions [2] rather than basing inferences only on non-pain opioid use disorder patients. Although opioid-induced neuroplastic changes are still poorly understood in humans, work has begun to illustrate these (briefly reviewed in [3]).

The amygdala is arguably the most relevant target due to its extensive reciprocal connections with pain and emotion processing and regulatory brain areas, its key roles in pain processing pathways, pain augmentation and negative affect circuit [4], pain progression [5, 6], hyperkatifeia^1^ [7], and the transition to substance addiction [8]. Moreover, the amygdala is the brain region with the densest mu opioid receptor (MOR) availability [9]. MOR have been shown in an animal study to be particularly relevant in opioid-induced hyperalgesia [10] and analgesic tolerance [11].

Opioid-related changes in amygdala connectivity was reported in opioid use disorder [12] but the reported hypoconnectivity may be specific to the underlying addiction. Opioid-related effects on amygdala network in chronic pain are not investigated but are likely given the link between MOR availability and migraine attacks frequency [13]. Interestingly, dominance of right amygdala relative to its left counterpart fits right amygdala dominance in pain progression [14], reported opioid-related volumetric changes [15], and even MOR availability in pain-free controls [16]; findings from a recent meta-analyses corroborate arguments on lateralisation when the left but not right amygdala was linked with positive emotions and mu opioid release [17].

The mediodorsal thalamic nuclei are similarly important regions for affective aspects of pain processing [18] and display a particularly high MOR density but also richness in kappa- and delta opioids receptors [19]. Indeed, a PET study identified the thalamus as the region with the highest opioid receptor density and demonstrated reduced thalamic opioid binding in response to experimental pain [20]. Interestingly, a left-sided predominance in thalamic opioid receptors has been suggested in direct opposition to the amygdala [16]. Such an opposite laterality was also found to be associated with chronicity of migraine as participants with chronic migraine demonstrated lower MOR availability in the left thalamus and the right amygdala relative to episodic migraineurs [13].

To address the knowledge gap of opioid-related alterations in these two networks in chronic pain we undertook a retrospective fMRI study on participants with chronic knee Osteoarthritis (OA) using opioids and matched pain participants without opioid prescription and compared right amygdala and left thalamic seed-whole brain functional connectivity maps between the two groups. We investigated resting state connectivity as inferences are not restricted to a specific paradigm and therefore have increased ecological validity that may be representative of effects on a person’s everyday life whilst also being more sensitive to changes that may not have manifested structurally yet. To explore associations between observed network changes and maladaptive symptoms, we undertook post-hoc correlations focused on psychometric and behavioural group differences.

## Methods

### Participants and inclusion criteria

This retrospective study used a subset of data from a multimodal pain phenotyping study that recruited participants with chronic knee Osteoarthritis (OA) pain, defined as pain for at least 3 months, and pain-free matched controls.

All participants gave informed consent and the study was approved by NRES Committee East Midlands - Nottingham 2 (REC reference number: 10/H0408/115) and performed in accordance with the declaration of Helsinki. Exclusion criteria at enrolment included contraindication to MRI or significant neurological, psychiatric or other significant health problem. Further information on the subject pool (whilst recruitment was ongoing) as well as data not reported here can be found in [21].

For the current analyses, participants were selected if they reported to take medical opioids for pain relief (KOPOp+). A control OA pain group (KOPOp-) not reporting to take any opioids was manually matched to age- and sex of the opioid use group on an individual basis and balanced at group level for pain burden (ICOAP) while being blinded about other characteristics of the data sets.

### Data for current analysis

MRI and phenotypic data relevant for the current analyses included quality controlled T1 and resting state fMRI data. Included questionnaires are summarised in table 1.

**Table 1:**
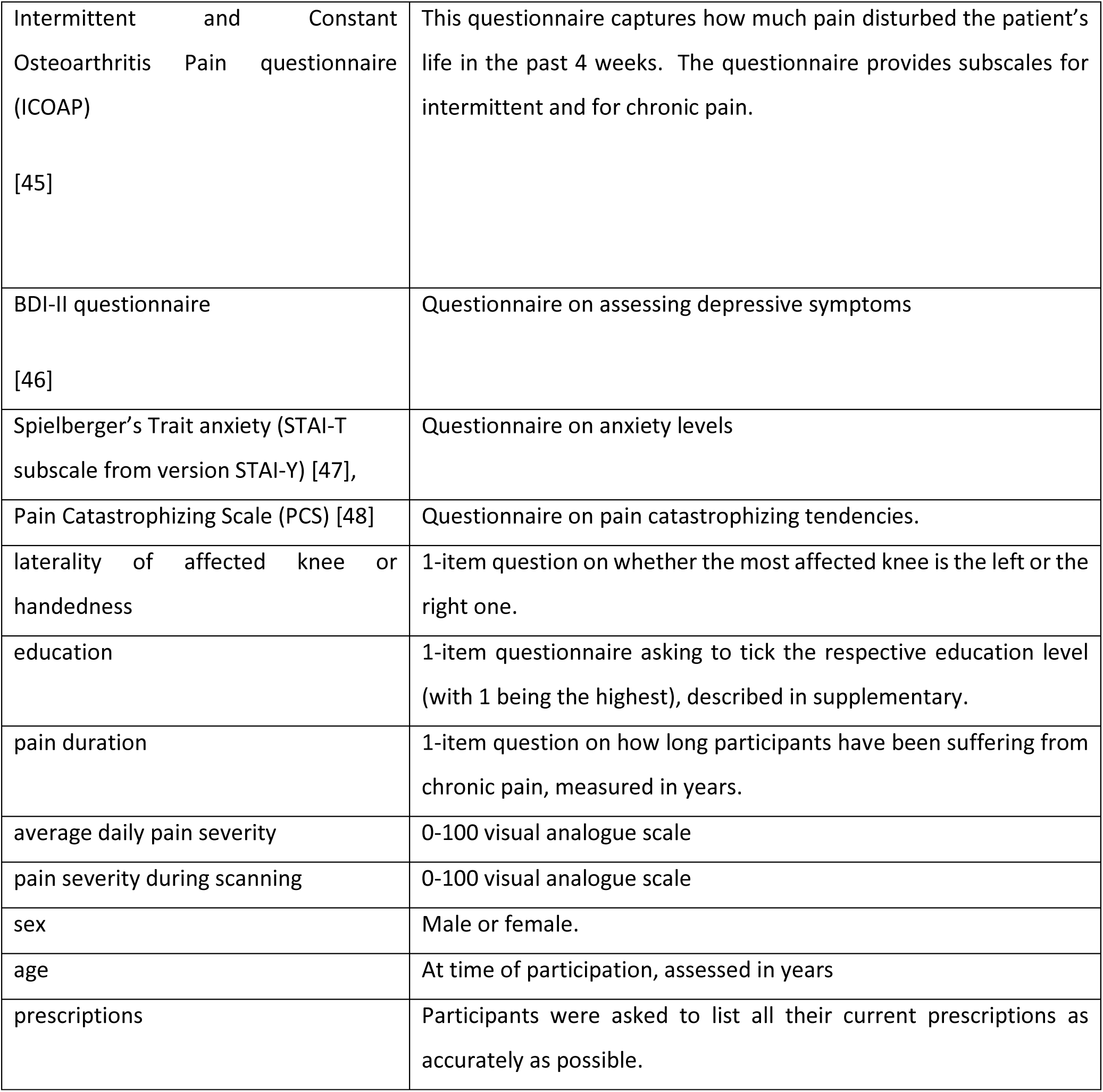
Questionnaires and demographic questions.

### Scan parameters

All participants were scanned at 3T (GE MR 750 Discovery, 32-channel head coil) following a multimodal protocol, with the current analysis focused on the standard anatomical (MPRAGE, TE/TR: 3.6/9.1s, flip angle: 12, sagittal acquisition, dimensions: 256×256×124, voxel size: 1mm^3^) and standard resting state functional scans using gradient echo echoplanar acquisition (200 volumes, TE/TR: 3.0/2.0s, voxel size: 3×3x 3.5mm, flip angle: 77, FOV 240, matrix 64×64×37, axial acquisition, phase encoding direction: A> P). Participants were asked to focus on a fixation cross in the middle of the screen, keep their eyes open and not to think of anything specific.

### Quality control and preprocessing

Preprocessing (brain extraction, motion correction with Eddy, slice timing correction, spatial smoothing with 5mm FWHM, ICA-AROMA for denoising, high-pass filtering) was performed with an in-house matlab script as described in [22]. MRI-QC software was used for quality control [23] with framewise displacement > 3mm or average framewise displacement >1mm as exclusion criteria.

Visual inspection of data was done at preprocessing individual and group analysis stage to check for any obvious coverage, signal drop-out or registration errors to MNI space.

All quality assurance was done blinded to participant characteristics of the respective data sets.

### Seed-based functional connectivity analysis

Analyses were performed with FSL feat version 6.0.1; all seeds were analysed separately.

#### Amygdala seeds

Harvard-Oxford amygdala masks at 80% probability thresholds were chosen (see supplementary) and registered to individual functional space to extract amygdala time series (left amygdala results are in supplementary).

#### Mediodorsal thalamic seeds

5mm diameter spheres around MNI coordinates x=8 y=-15 z=6 and x=-6 y=-15 z=7 depicting the centre of gravity of the mediodorsal thalamic nuclei according to WFU Pick Atlas v3.0.5 were used as thalamic seed regions (figure in supplementary). Only left thalamus FC is reported here (results for the right counterpart are in supplementary).

### Network reconstruction

FSL’s FAST was used to segment T1-weighted images into cerebrospinal fluid (CSF) and white matter (WM) which were then thresholded (volume x probability) to avoid partial volume effects using an in-house matlab script [22], registered to individual functional space via FSL’s FLIRT, and used as masks to extract CSF and WM time series for each data set (using fslmeants) from the denoised functional images (for regressors of no interest).

First level analysis of the time-series was performed on the denoised data in native space separately for each seed’s time series. For this, amygdala masks were non-linearly registered to subject space and time series extraction was performed with fslmaths. The extracted time series was inserted as a variable of interest in FSL’s FEAT while correcting for CSF and WM as covariates of no interest using the time series extracted as described above. This step further used FILM in FSL’s FEAT for autocorrelation correction (prewhitening) [24].

### Group comparisons and statistical inference

Independent samples t-tests were performed to check for demographic or pain-related group differences.

To compare fc networks of each seed region between opioid prescribed and non-prescribed participants, a general linear model was used. The fsl flameo command was performed on the 4D file containing all subjects after individual network maps from first level analysis were non-linearly registered to MNI template. The design matrix was set up via FSL and included demeaned relative motion, sex, and age, per person as covariates of no interest. A gray matter mask was used to limit multiple comparison correction to gray matter voxels only. Cluster-based p-value threshold was set at 0.05, z-values >2.3 to enable comparisons with published reports typically still based on this threshold. In addition, we report results at the more stringent recently recommended z-value >3.1 threshold; significant clusters for both z thresholds are overlaid in figure 1.

**Figure 1:**
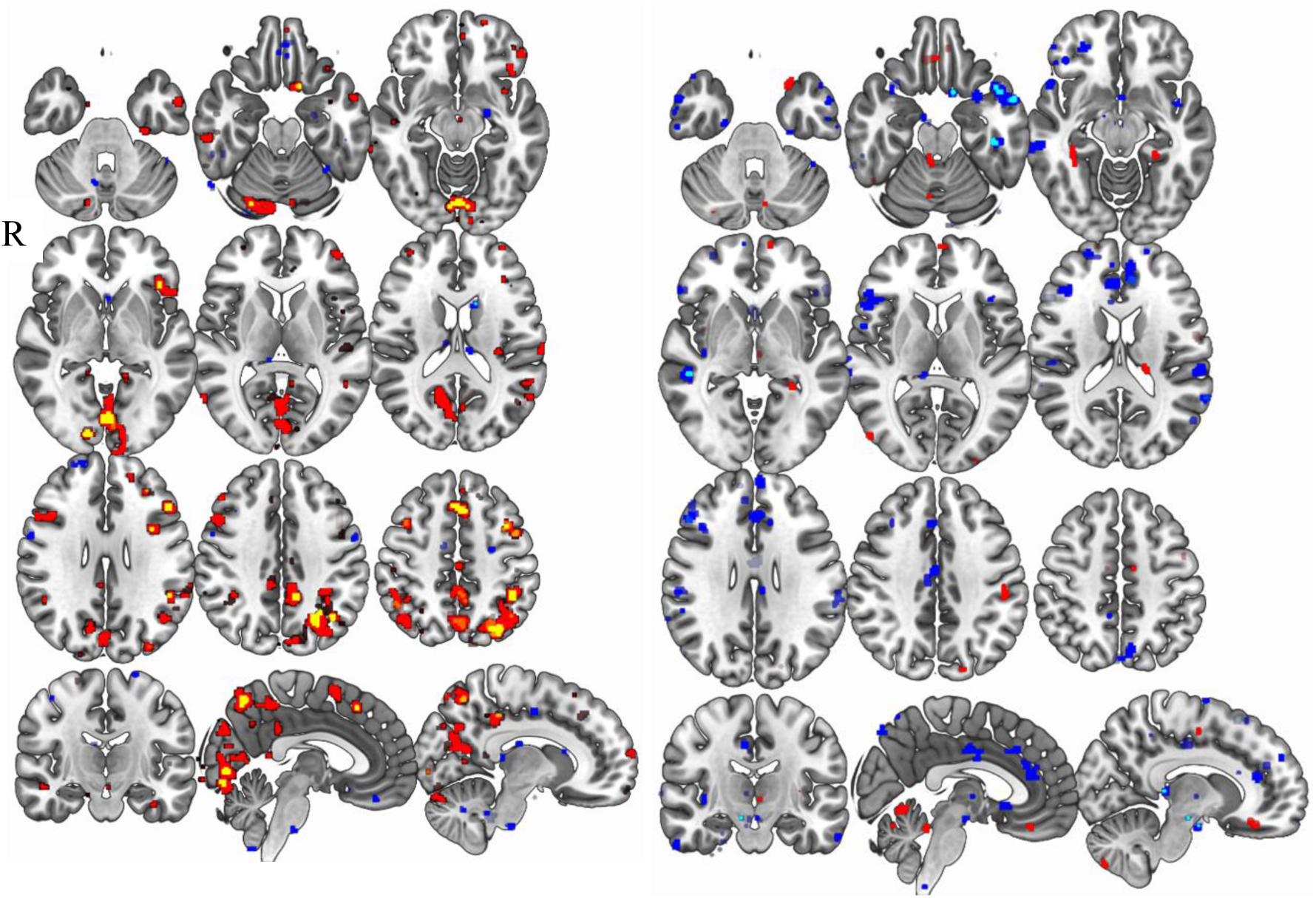
Group differences in right amygdala and left mediodorsal thalamic FC networks. The left side of figure 1 shows the results for the right amygdala seed region while the other part of the figure shows results for the left thalamus seed. The figure shows the contrast Opioid prescribed knee pain participants > control participants: red (z-score min. 2.3)-yellow (z-score min. 3.1) and Control participants > opioid prescribed participants: blue (z-score min. 2.3)- lightblue (z-score min. 3.1).

Within-group maps are provided in the supplementary for reference.

### Post-hoc tests

We visually compared within-group FC maps for amygdala and thalamic networks to discern whether significant group differences indicate increased positive or decreased negative functional connectivity.

To explore whether FC changes may be related to negative affect, we extracted z-scores from most prominent clusters and undertook Pearson correlations with trait anxiety scores across both groups. Scatterplots of these correlations are in the supplementary.

## Results

### Demographic and pain-related data

Fourteen participants (8 males), reporting to take prescription opioids, had high quality FC data and were matched with 13 participants (8 males) as opioid-free knee OA control group (KOPOp-) (table 2). Apart from depressive symptoms and trait anxiety levels, characteristics did not significantly differ between the groups (table 2).

**Table 2:**
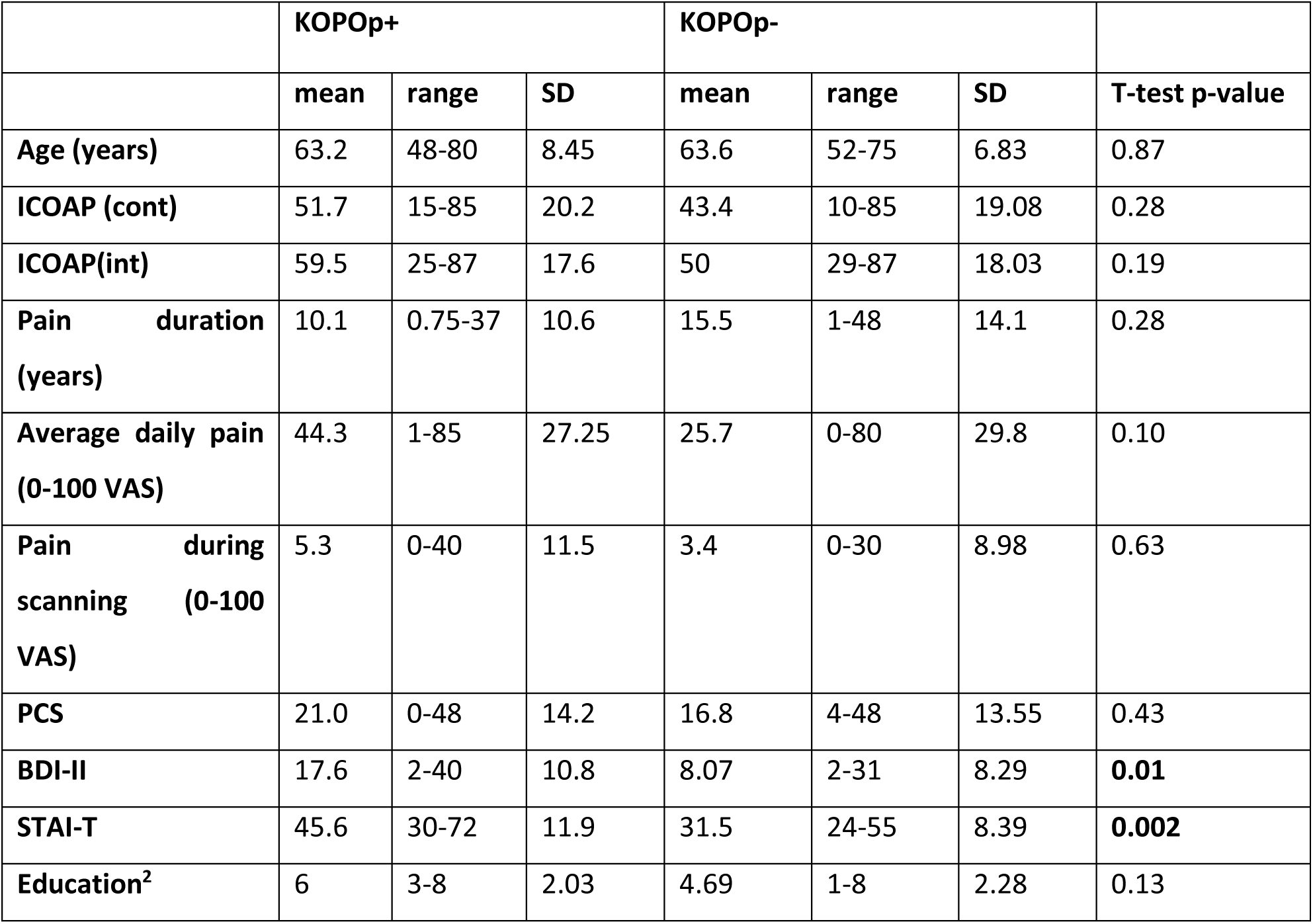
Characteristics of opioid (KOPOp+) and non-opioid using pain participant (KOPOp-) groups.

|  | KOPOp+ |  |  | KOPOp- |  |  |  |
| --- | --- | --- | --- | --- | --- | --- | --- |
|  | mean | range | SD | mean | range | SD | T-test p-value |
| Age (years) | 63.2 | 48-80 | 8.45 | 63.6 | 52-75 | 6.83 | 0.87 |
| ICOAP (cont) | 51.7 | 15-85 | 20.2 | 43.4 | 10-85 | 19.08 | 0.28 |
| ICOAP(int) | 59.5 | 25-87 | 17.6 | 50 | 29-87 | 18.03 | 0.19 |
| Pain duration (years) | 10.1 | 0.75-37 | 10.6 | 15.5 | 1-48 | 14.1 | 0.28 |
| Average daily pain (0-100 VAS) | 44.3 | 1-85 | 27.25 | 25.7 | 0-80 | 29.8 | 0.10 |
| Pain during scanning (0-100 VAS) | 5.3 | 0-40 | 11.5 | 3.4 | 0-30 | 8.98 | 0.63 |
| PCS | 21.0 | 0-48 | 14.2 | 16.8 | 4-48 | 13.55 | 0.43 |
| BDI-II | 17.6 | 2-40 | 10.8 | 8.07 | 2-31 | 8.29 | **0.01** |
| STAI-T | 45.6 | 30-72 | 11.9 | 31.5 | 24-55 | 8.39 | **0.002** |
| Education<sup>2</sup> | 6 | 3-8 | 2.03 | 4.69 | 1-8 | 2.28 | 0.13 |

<sup>2</sup> Level ranges from 8: none to 1: higher degree, postgraduate degree. See supplementary for more details.

### Functional connectivity results: Amygdala FC

Relative hyperconnectivity of the right amygdala functional network was seen in participants with chronic knee OA pain reporting to take opioids relative to their matched control pain participants. The network changes included occipital areas, the precuneus and other parts of the default mode network (DMN), fronto-parietal areas and the dorsomedial prefrontal cortex (dmPFC) (Figure 1, table 3). Inspection of within-group network maps as post-hoc tests demonstrated that clusters are anti-correlated with the right amygdala in KOPOp-but are positively correlated in KOPOp+ (see supplementary), indicating a reversal of coupling characteristics with prolonged prescription opioid use. The opposite contrast (relative hypoconnectivity changes in KOPOp+) did not detect clusters at the chosen cluster extent threshold (table 3 but see figure 1 without minimum cluster size). To explore interrelationship with higher negative affect scores in prescription opioid users, further post-hoc tests showed a significant positive correlation between right amygdala-posterior DMN connectivity and trait anxiety scores (p=0.001, r=0.61).

**Table 3:** Network FC group differences for clusters with > 20 voxels.

| **Seed: Right Amygdala** |  |  |  |  |  |
| --- | --- | --- | --- | --- | --- |
| **<u>Opioid participants > Control participants</u>** | Voxel count | MAX Z | MAX X (vox) | MAX Y (vox) | MAX Z (vox) |
| Left Lateral Occipital Cortex | 947 | 4.57 | 58 | 32 | 55 |
| Left Lingual Gyrus | 746 | 3.78 | 46 | 24 | 31 |
| Right Precuneus | 400 | 3.61 | 43 | 31 | 63 |
| Left Precuneus | 306 | 3.38 | 50 | 39 | 56 |
| Left Middle Frontal Gyrus | 276 | 4.49 | 64 | 68 | 65 |
| Right Lateral Occipital Cortex | 237 | 3.51 | 22 | 36 | 62 |
| Right Cuneus | 174 | 3.04 | 40 | 29 | 45 |
| Superior Frontal Gyrus/dmPFC | 171 | 3.38 | 46 | 74 | 60 |
| Left Middle Frontal Gyrus | 167 | 3.5 | 69 | 74 | 49 |
| Left Frontal Orbital Cortex | 130 | 3.32 | 65 | 77 | 33 |
| Left Precuneus | 127 | 3.51 | 48 | 33 | 42 |
| Right Middle Frontal Gyrus | 92 | 3.24 | 20 | 70 | 56 |
| Cerebellum | 68 | 3.58 | 53 | 40 | 7 |
| Cuneus | 62 | 2.98 | 44 | 24 | 51 |
| Left Superior Frontal Gyrus | 61 | 3.08 | 49 | 77 | 67 |
| Left Supramarginal Gyrus | 50 | 3.15 | 70 | 40 | 50 |
| Right Middle Frontal Gyrus | 45 | 3.4 | 25 | 69 | 62 |
| Left Frontal Pole | 40 | 3.03 | 66 | 89 | 42 |
| Right Occipital Pole | 39 | 3.74 | 38 | 18 | 35 |
| Left Posterior Cingulate | 37 | 3 | 51 | 52 | 58 |
| Left Lateral Occipital Cortex | 32 | 3.21 | 62 | 20 | 51 |
| Right Inferior Temporal Gyrus | 32 | 2.93 | 16 | 49 | 25 |
| Posterior Cingulate | 29 | 2.92 | 42 | 41 | 41 |
| Left Frontal Orbital Cortex | 23 | 3.24 | 52 | 69 | 25 |
| Right Lateral Occipital Cortex | 22 | 2.68 | 31 | 33 | 66 |
| Right Middle Frontal Gyrus | 22 | 3.42 | 21 | 78 | 54 |
| **<u>Control participants > Opioid participants</u>** |  |  |  |  |  |
| No cluster with min. 20 voxels. |  |  |  |  |  |

Prescription-opioid amygdalar & thalamic network alterations
| **Seed: Left thalamus** |  |  |  |  |  |
| --- | --- | --- | --- | --- | --- |
| **<u>Opioid participants > Control participants</u>** | Voxel count | MAX Z | MAX X (vox) | MAX Y (vox) | MAX Z (vox) |
| Left Hippocampus | 35 | 2.69 | 57 | 44 | 33 |
| Left Supramarginal Gyrus | 29 | 3.55 | 71 | 50 | 53 |
| Cerebellum | 25 | 2.62 | 43 | 32 | 32 |
| Left Frontal Orbital Cortex | 25 | 3.32 | 58 | 72 | 22 |
| Right vmPFC | 22 | 2.78 | 41 | 80 | 26 |
| **<u>Control participants > Opioid participants</u>** |  |  |  |  |  |
| Left Temporal Pole | 174 | 3.44 | 73 | 65 | 24 |
| Anterior Cingulate | 133 | 3.78 | 50 | 83 | 43 |
| Right Middle Frontal Gyrus | 105 | 3.29 | 20 | 77 | 51 |
| Posterior Cingulate | 68 | 2.99 | 41 | 54 | 56 |
| Right Inferior Frontal Gyrus | 61 | 2.87 | 18 | 75 | 43 |
| Right Superior Frontal Gyrus | 59 | 4.09 | 37 | 74 | 67 |
| Left Supramarginal Gyrus | 56 | 2.75 | 76 | 47 | 48 |
| Right Middle Temporal Gyrus | 45 | 3.18 | 20 | 47 | 35 |
| Right Frontal Gyrus | 40 | 2.88 | 37 | 91 | 46 |
| Right Middle Temporal Gyrus | 40 | 3.05 | 10 | 48 | 30 |
| Right Supramarginal Gyrus | 37 | 3.22 | 14 | 44 | 42 |
| Cerebellum | 35 | 3.01 | 34 | 29 | 8 |
| Right Middle Temporal Gyrus | 34 | 2.88 | 15 | 61 | 22 |
| Left Supramarginal Gyrus | 33 | 3.42 | 78 | 38 | 44 |
| Left Precuneus | 30 | 2.75 | 49 | 28 | 60 |
| Subgenual Anterior Cingulate | 28 | 3.41 | 46 | 70 | 33 |
| Anterior Cingulate | 27 | 3.01 | 46 | 78 | 50 |
| Right Supramarginal Gyrus | 26 | 2.84 | 17 | 50 | 49 |
| Cerebellum | 24 | 3.34 | 33 | 42 | 14 |
| Cerebellum | 20 | 3.7 | 24 | 34 | 11 |
| dmPFC | 20 | 2.93 | 47 | 92 | 50 |

The same directionality was found for the amygdala-dmPFC group difference (Figure 1, table 3 and supplementary material for within-group FC maps) but there was no correlation with trait anxiety.

Left amygdala network differences between the two groups were beyond the scope of the current study but show similar findings and are included in the supplementary material for completeness.

### Dorsomedial thalamic FC

In the left thalamic network on the other hand, there was predominant hypoconnectivity with cingulate, frontolateral and temporal areas (esp. left temporopolar) and right supramarginal cortex in opioid users relative to controls (table3, figure 1) whilst there were some foci of hyperconnectivity, such as with the hippocampus. Comparison with within-group FC maps as post-hoc tests reveals that cingulate clusters denote positive connectivity for both groups, confirming less positive FC for these regions with the mediodorsal thalamic nucleus in the KOPOp+ group with the notable exception of the subgenual anterior cingulate cluster which was anti-correlated with the seed region in KOPOp+. The latter clusters were absent in controls (see supplementary material). Supplementary material also includes group differences in right mediodorsal thalamic network with visually similar patterns.

No tendency for correlations between mediodorsal thalamic-mid cingulate and mediodorsal thalamic-anterior cingulate FC and trait anxiety was found.

## Discussion

Using seed-based functional connectivity analysis in participants with chronic painful knee OA, we report distinct brain network changes of MOR rich hubs of the affective pain processing in association with opioid prescription status. The main right amygdala fc change associated with opioid status was a distributed pattern of hyperconnectivity involving the posterior DMN, the visual cortex, left supramarginal gyrus and dorso-medial prefrontal cortex significant at z >3.1. Medio-dorsal thalamic networks changes related to opioid prescription consisted of a predominant hypoconnectivity pattern with several cingulate regions and the left anterior temporal pole, were detected at Z>2.3, but did not survive the Z> 3.1 threshold. We chose to report results at both thresholds to allow comparisons across most published reports because the switch to a more conservative Z>3.1 is still relatively recent and because the small sample size of this discovery study intends to inform further work which justifies a more inclusive approach.

We found stronger positive coupling between the right amygdala and posterior DMN (pDMN) in opioid prescribed knee OA participants compared to controls whilst controls showed an anti-correlation between the right amygdala and the pDMN. Contrary to this study in opioid using participants, however, a previous FC study in participants with opioid addiction reported reduced amygdala functional connectivity with the pDMN [12]. The cause of that discrepancy is unclear but may be due differences in the type and cumulative dose of opioids and study populations (opioid addiction versus prescription opioids for analgesia in the present study) and different comparison groups (healthy controls versus matched pain participants in the present study). Interestingly, acute oxycodone exposure in animals [25] and humans [26] was reported to decrease DMN FC including the amygdala which suggests that the observed opioid-related amygdala-DMN FC hyperconnectivity in our study does not simply reflect acute opioid effects and more likely represents a marker of network plasticity. We also found that the between group FC hyperconnectivity was driven by a remarkable directionality shift from anticorrelation in non-opioid users to positive coupling in opioid users. There are two main possible causes for this circuit abnormality either as pre-existing endophenotypic trait predisposing the need for stronger pain killers or a consequence of such, which we cannot discern in our cross-sectional study.

We observed a moderately strong positive correlation across both groups between the right amygdala–pDMN FC and trait anxiety making a maladaptive nature of the circuit change more likely. This is well in line with reports on increased FC of the amygdala-pDMN in affective disorders compared to healthy populations. Enhanced FC in the amygdala-pDMN circuit was shown in major depression even after remission, and FC changes were linked to high levels of rumination [27]. Similarly, increased amygdala-pDMN FC was linked to late life depression [28]. Stress alone may increase amygdala-pDMN FC [29] as shown in a stress induction task in healthy controls providing a relevant context for interpretation in chronic pain beyond depression.

Indeed, higher FC between the pDMN (labelled as ventral DMN by the authors) and parts of the amygdala was linked to less resilience but irrespective of depression [30], which would suggest that our observation may be linked more generally to decreased resilience in opioid using pain participants as additional maladaptive mechanisms beyond dispositional anxiety and negative affect. Further complexity may arise from frequent co-administration of antidepressants in the opioid using pain group as antidepressant infusion reversed a positive correlation between amygdala and DMN in depressed patients [31]. This makes antidepressant co-medication highly unlikely to have contributed to our observed pattern of increased amygdala-pDMN FC.

We observed a noteworthy amygdala-dmPFC hyperconnectivity in opioid using compared to opioid-free pain participants and a similar shift from an anti-correlated relationship between these two brain structures in opioid-free controls. The dmPFC is considered an ‘aversive amplification’ circuit [32], that underlies ‘negative bias’ [33], and may even enable maintenance of anxiety in healthy volunteers [34] and the development of clinical anxiety [35] which suggests a link with anxiety even if some of the labelling should be taken with caution; however our post-hoc tests showed no tendency for a correlation between this connection and trait anxiety. A shift towards synchronization between the amygdala and the dmPFC from prolonged opioid use may therefore contribute to maladaptive behavioural developments through a more complex mechanism. Interestingly, serotonin depletion, an experimental model of depression, was linked with increased amygdala-dmPFC FC in healthy volunteers [33] while antidepressant treatment and symptom improvement were linked to decreased dmPFC FC [36]. This again suggests that common co-medication with antidepressants in opioid prescribed chronic pain participants is unlikely to drive the observed hyperconnectivity of this circuit in opioid users.

Conversely, we found predominant hypoconnectivity of the mediodorsal thalamo-cortical network in association with opioid prescription status involving several clusters in the cingulum.

Similar to the thalamus and the amygdala, the cingulate is tightly linked to affective processing in pain. Specifically, posterior parts of the anterior cingulate have been implicated with unpleasantness, anterior parts of the mid cingulate (ACC) have been implicated with processing fear whilst subgenual parts are linked with negative stimuli [37] and play a prominent role in depression [38]; ACC hyperactivity has even been found to be characteristic of pain-induced depression and anxiety [39, 40] but our post-hoc correlations showed no tendency for a simple link with trait anxiety.

More generally, animal literature suggested that anterior cingulate hyperactivity plays a key role in pain progression [40-42] but how this hyperactivity relates to thalamic-cingulate connectivity has not been investigated in humans.

An animal study, however, showed that neurons within the ACC responded less strongly to mediodorsal thalamic input as a consequence of induced chronic pain such that the ACC was inhibited which in turn was shown to be directly enhancing pain-related aversion [43]. In the context of the current findings it would suggest that prolonged use of exogenous opioids exacerbates pain-related aversion through mediodorsal thalamic-cingulate hypoconnectivity; this in turn could reflect either maladaptive circuit alterations or impaired compensatory mechanisms. Against the background of Zubieta et al.’s finding that more mu-opioid receptor binding in the cingulate was linked with less negative affect in sad settings [44], one may speculate that the cingulum is normally part of a pain compensatory circuit which appears to be altered after prolonged opioid use. This could potentially reflect receptor fatigue but remains to be studied further.

The main limitations of the current work are the small sample size and its retrospective and cross-sectional design which makes it impossible to disentangle opioid-related from co-medicated antidepressants or pre-existing circuit abnormalities biasing towards the need for opioidergic pain killers. Further, due to the retrospective nature of this study we lack information on opioid use duration or any opioid induced hyperalgesia or tolerance.

Overall however, the current work shows that resting state amygdalar and thalamic networks are differentially altered in chronic pain participants on prolonged opioid use compared to opioid-free participants suggestive of opioid-related maladaptive developments.

## Data Availability

Data used in this study may be available upon reasonable request.

## Author contributions

DPA provided the concept for this study, DPA and MMD contributed the design and analysis plan; MMD analysed the data with support by WJC, SJI and AMN; data interpretation and manuscript drafting was done by MMD and DPA; WJC, SJI, MMD, AT collected data. All co-authors were involved in editing and proof-reading.

## Acknowledgements

We thank all participants who kindly volunteered their time to this study. We are also thankful to Nadia Frowd and Dr Bonnie Millar for administrative support and recruitment, Andrew Cooper for scanning with us as radiographer, Sarah Wilson as the centre’s dedicated receptionist and admin, and to the KPIC data collection team for helping our recruitment efforts by sharing contact details of participants who consented to be contacted for further studies and met eligibility for the current one. We additionally thank Prof. D. Walsh for comments.

## Footnotes

1 withdrawal induced hypersensitivity to negative emotions

2 Level ranges from 8: none to 1: higher degree, postgraduate degree. See supplementary for more details.

