## supplementary for "Prescription opioid-related alterations to amygdalar and thalamic functional networks in chronic knee pain: A retrospective case control resting-state connectivity study"

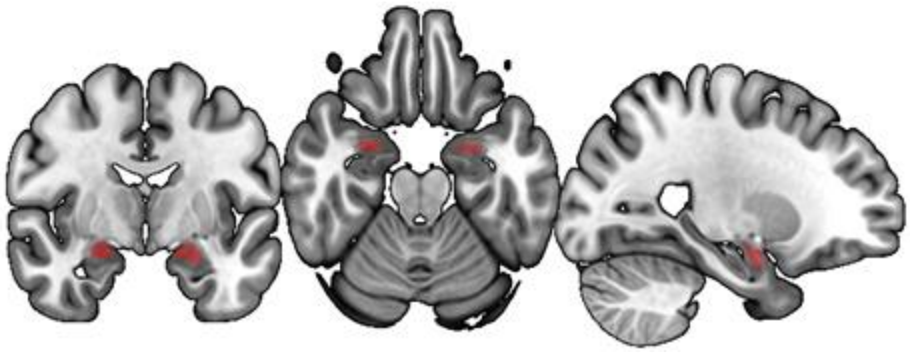

*Figure 1: Amygdala masks*

*Figure 1 shows the masks used in the current analyses which were based on amygdala masks from the Harvard-Oxford probabilistic atlas and thresholded at 80% probability for a voxel to be part of the amygdala. Only the right amygdala seed was part of the main results.*

This atlas was chosen based on prior visual checks confirming adequate anatomical accuracy of these masks; the 80% threshold was chosen over the commonly applied 50% threshold to minimize inclusion of the hippocampi and minimize further anatomical inaccuracy after smoothing. The volume of right amygdala seed mask was 114 voxels.

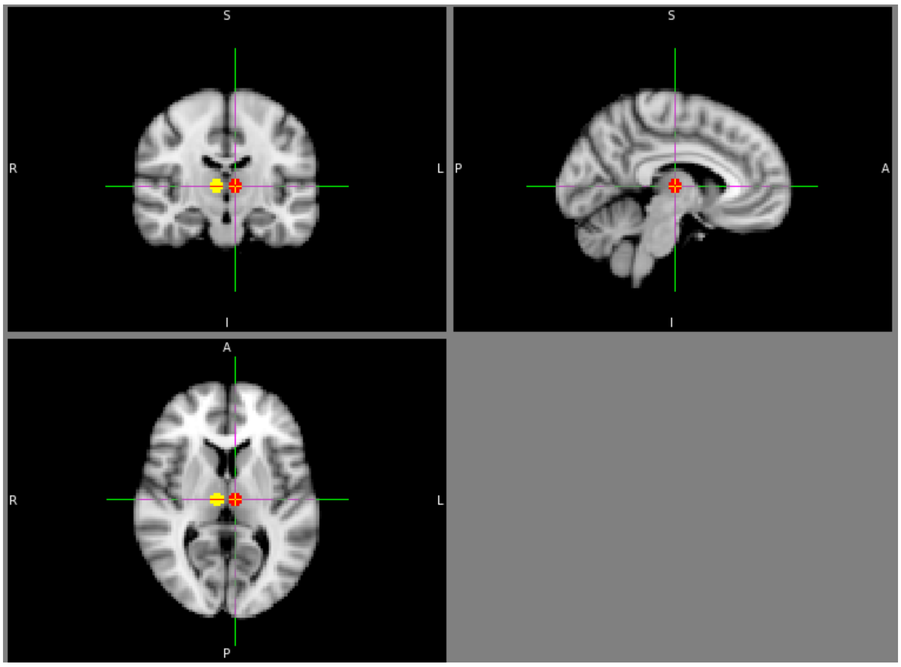


*Figure 2: Thalamic mask*

*Figure 1 shows the masks used in the current analyses; note that the masks were analysed separately and only the left thalamus seed was part of the main results.*

The volume of the left mediothalamic nucleus mask encompassed 81 voxels.

### Supplementary

*Subject pool*

The overall data set from the multimodal study contained 120 subjects at the time of analysing. N=80 out of n=120 were pain participants with chronic knee OA. Exclusion of data sets was done blinded to grouping: n=8 for not passing quality assurance, n=1 for scanner artefact, n=1 for considerable dropout, n=1 for incidental finding.

Of the remaining pain participant data sets, n=14 pain participants were identified with self-reported opioid prescriptions; 13 control pain participants were identified that could be matched to the sample as described in the methods.

*Education scores:*

*Participants were asked to indicate their education level on this form:*

*Please indicate the answer that best describes*
*your educational attainment:*

1 = Higher degree and professional qualifications, Postgraduate degree/qualifications
2 = Undergraduate degree
3 = Diploma, certificate (tertiary qualification below degree level)
4 = Other sub-degree (tech. & business qualifications above A-level but below degree level)
5 = A and AS level GCE
6 = Below AS level (0-level GCE/ GCSE, CSE, vocational qual. below sub-degree level)
7 = Apprenticeship
8 = None

**Table 1: Self-reported usage of prescription opioids, non-opioid pain killers, and antidepressants**

| Participant | Opioids | Non-opioid pain killers | Antidepressants |
|---|---|---|---|
| 1 | Codeine phosphate | Voltarol gel<br>Paracetamol | Amitriptyline |
| 2 | Tramadol | Paracetamol<br>Rizatriptan (anti-migraine) | Amitriptyline<br>Citalopram |
| 3 | Cocodamol<br>Codeine | None reported | None reported |
| 4 | Zapain (Codeine Phosphate/Paracetamol) | Aspirin | Citalopram |
| 5 | Co-codamol | Fenbid Forte gel, Naproxen, Nefopam | Amitriptyline |
| 6 | Tramadol,<br>Dihydrocodeine (20mg) | Ibuprofen (400mg),<br>Paracetamol | None reported |
| 7 | Morphine (patches) | Naproxen | Seroxin (100mg),<br>Amitriptyline |
| 8 | Codeine phosphate (15mg) | Naproxen(500mg),<br>Paracetamol(500mg) | None reported |
| 9 | Zapain (codeine) (30mg) | Paracetamol(500mg), Fenbid (Ibuprofen) | None reported |
| 10 | Dihydrocodeine (240mg) | None reported | Amitriptyline (75mg) |

| | | | |
|---|---|---|---|
| 11 | Co-codamol | None reported | None reported |
| 12 | Dihydrocodeine (30mg) | Naproxen (500mg), Paracetamol | Amitriptylin (25mg) |
| 13 | Co-codamol (8mg) | Fenbid gel (Ibuprofen) | Sertraline (100 mg) |
| 14 | Tramadol (2mg) | Aspirin (75 mg), Paracetamol (500 mg) | None reported |
| Control pain participants | | | |
| 15 | None reported | Ibuprofen, Paracetamol | None reported |
| 16 | None reported | None reported | None reported |
| 17 | None reported | None reported | None reported |
| 18 | None reported | None reported | None reported |
| 19 | None reported | None reported | None reported |
| 20 | None reported | None reported | None reported |
| 21 | None reported | Paracetamol, Aspirin | None reported |
| 22 | None reported | None reported | None reported |
| 23 | None reported | Naproxen (500mg) | None reported |
| 24 | None reported | None reported | None reported |
| 25 | None reported | Paracetamol, Ibuprofen, Aspirin (75mg) | None reported |
| 26 | None reported | None reported | None reported |
| 27 | None reported | Naproxen (1000mg/d) | None reported |

**Analysis**
**Network reconstruction**

FSL's FAST was used to segment T1-weighted images into cerebrospinal fluid (CSF) and white matter (WM) which were then thresholded (volume x probability) to avoid partial volume effects using an in-house matlab script (Mohammadi-Nejad et al.), registered to individual functional space via FSL's FLIRT, and used as masks to extract CSF and WM time series for each data set (using fslmeants) from the denoised functional images (for regressors of no interest).

**Within-group FC maps**

These maps represent the within group functional connectivity maps.

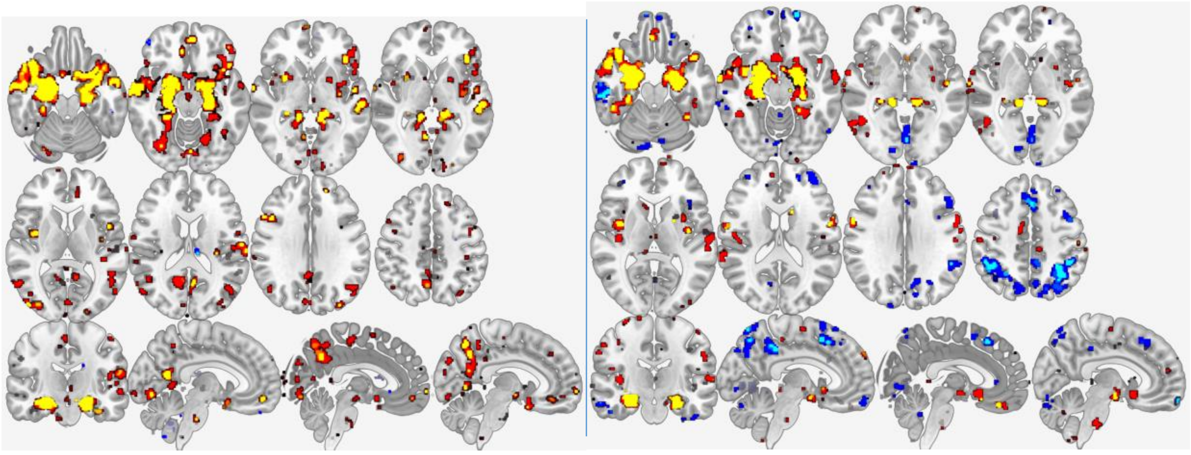

**Figure 3: Right Amygdala**
Left: opioid prescribed pain participants, Right: control pain participants.
Red (z-score min.2.3) –yellow (z-score min.3.1) positive connectivity,
blue (z-score min.2.3) –lightblue (z-score min.3.1): negative connectivity.
Radiological convention.

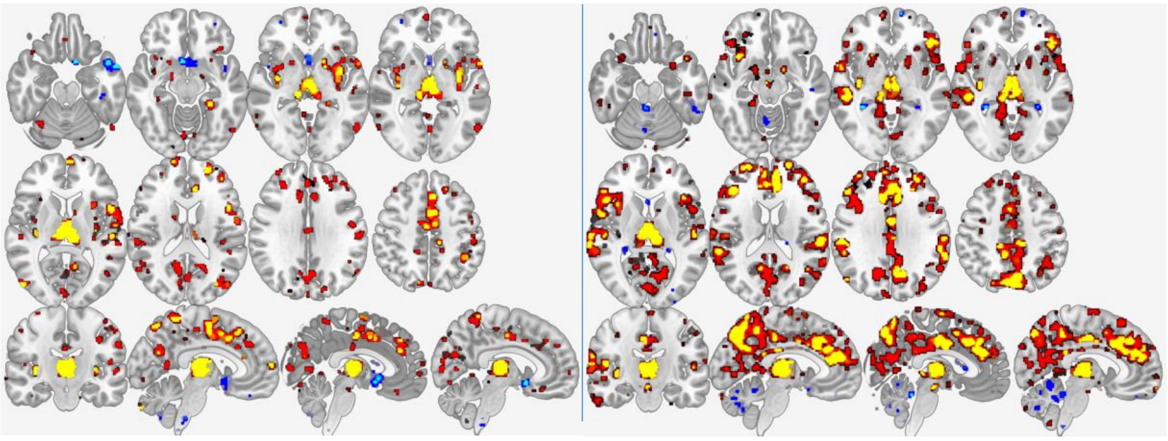

**Figure 4: Left thalamus seed**
Left: opioid prescribed pain participants, Right: control pain participants.
Red (z-score min.2.3) –yellow (z-score min.3.1) positive connectivity,
blue (z-score min.2.3) –lightblue (z-score min.3.1): negative connectivity.
Radiological convention.

**Results from contralateral seed not shown in results section:**

Table 2: Amygdala FC

| LEFT AMYGDALA | | | | | |
|---|---|---|---|---|---|
| **Opioid participants > Control participants** | Voxel count | MAX Z | MAX X (vox) | MAX Y (vox) | MAX Z (vox) |
| Left supramarginal gyrus/planum temporale | 150 | 3.47 | 72 | 39 | 44 |
| Left Occipital Pole | 103 | 3.81 | 49 | 14 | 32 |
| Right Precuneus | 91 | 3.62 | 44 | 42 | 61 |
| Right Occipital Cortex | 88 | 3.24 | 39 | 18 | 34 |
| Left Middle Frontal Gyrus | 82 | 3.04 | 65 | 64 | 61 |
| Left Insula | 69 | 3.35 | 64 | 54 | 41 |
| Right Lingual Gyrus | 68 | 3.87 | 39 | 43 | 33 |
| Cerebellum | 60 | 3.33 | 39 | 33 | 10 |
| Left inferior frontal gyrus | 60 | 3.21 | 68 | 80 | 36 |
| Right Cuneus | 50 | 2.82 | 41 | 26 | 47 |
| Left Planum Temporale | 40 | 2.95 | 70 | 53 | 39 |
| Left Occipital Cortex | 36 | 3.06 | 58 | 30 | 52 |
| Right Frontal Pole | 35 | 2.93 | 42 | 96 | 30 |
| Left Occipital Pole | 32 | 3.51 | 55 | 17 | 28 |
| Right Precuneus | 32 | 2.98 | 36 | 31 | 52 |
| Right vmPFC | 32 | 3.67 | 40 | 88 | 33 |
| Cerebellum | 31 | 2.88 | 55 | 40 | 9 |
| Left Temporal Fusiform Gyrus | 30 | 3.23 | 62 | 48 | 23 |
| Right Middle Temporal Gyrus | 25 | 2.7 | 18 | 62 | 23 |
| Left Superior Frontal Gyrus | 25 | 2.95 | 48 | 73 | 61 |
| Left Middle Frontal Gyrus | 24 | 3.11 | 66 | 70 | 53 |
| Right Occipital Cortex | 23 | 3.01 | 30 | 32 | 65 |
| Left Middle Temporal Gyrus | 20 | 2.75 | 69 | 59 | 27 |
| Right Thalamus | 20 | 3.21 | 42 | 54 | 35 |
| Right Temporal Pole | 20 | 2.66 | 21 | 70 | 22 |
| **Control participants > Opioid participants** | | | | | |
| RVM*[1] | 71 | 4.17 | 43 | 48 | 14 |
| Right Precentral Gyrus | 31 | 3.28 | 24 | 59 | 67 |
| Right Precentral Gyrus | 24 | 2.98 | 31 | 57 | 64 |
| Right Precentral Gyrus | 23 | 3.86 | 17 | 66 | 49 |
| Cerebellum | 22 | 3.41 | 43 | 36 | 10 |

[1] Some have discussed a cluster similar to the RVM in the current work as parabrachial nucleus (see figure 2 in review by 47. Bushnell, M.C., M. Ceko, and L.A. Low, *Cognitive and emotional control of pain and its disruption in chronic pain.* Nature Reviews Neuroscience, 2013. **14**(7): p. 502-511.

| | | | | | |
|---|---|---|---|---|---|
| Left Thalamus | 21 | 3.31 | 45 | 58 | 40 |

Table3: Right Thalamus FC

| Right thalamus | | | | | |
|---|---|---|---|---|---|
| **Opioid participants > Control participants** | Voxel count | MAX Z | MAX X (vox) | MAX Y (vox) | MAX Z (vox) |
| Cerebellum | 41 | 3.51 | 65 | 25 | 19 |
| Cerebellum | 41 | 3.86 | 50 | 27 | 16 |
| Left Postcentral gyrus | 39 | 3.16 | 70 | 51 | 57 |
| Right Postcentral gyrus | 35 | 3.22 | 37 | 40 | 70 |
| Cerebellum | 31 | 3.14 | 41 | 26 | 16 |
| Cerebellum | 27 | 3.13 | 25 | 26 | 21 |
| Left Superior Frontal Gyrus | 27 | 2.92 | 50 | 62 | 73 |
| Cerebellum | 25 | 3.19 | 30 | 23 | 15 |
| Right Caudate | 22 | 3.24 | 37 | 64 | 45 |
| Mid Cingulate | 21 | 3.62 | 50 | 59 | 58 |
| **Control participants > Opioid participants** | | | | | |
| Left insula | 401 | 3.88 | 65 | 64 | 29 |
| Right Middle Frontal Gyrus | 205 | 3.46 | 21 | 78 | 48 |
| Right Temporal Pole/insula | 125 | 3.24 | 21 | 72 | 29 |
| Right Angular Gyrus | 120 | 3.24 | 12 | 40 | 49 |
| Left Planum Polare/insula | 92 | 3.55 | 71 | 57 | 37 |
| Right Middle Temporal Gyrus | 79 | 3.17 | 14 | 59 | 28 |
| Right Temporal Pole/insula | 78 | 3.22 | 24 | 66 | 37 |
| Right Middle Temporal Gyrus | 72 | 3.19 | 13 | 58 | 23 |
| Left Temporal Pole | 69 | 3.35 | 74 | 66 | 23 |
| Left Precuneus | 67 | 2.84 | 48 | 29 | 57 |
| Right Parahippocampus | 54 | 3.46 | 36 | 51 | 24 |
| Right Superior Frontal Gyrus | 49 | 2.95 | 34 | 72 | 64 |
| Left Middle Temporal Gyrus | 39 | 3.52 | 79 | 51 | 31 |
| Right Hippocampus | 37 | 2.96 | 40 | 59 | 26 |
| Anterior Cingulate | 36 | 3.26 | 46 | 81 | 43 |
| Brainstem | 36 | 3.61 | 47 | 56 | 28 |
| Right Temporal Pole | 29 | 3.14 | 16 | 66 | 20 |
| Right Frontal Pole | 28 | 2.96 | 22 | 80 | 40 |
| Left Middle Temporal Gyrus | 25 | 3.01 | 68 | 33 | 42 |
| Right Frontal Orbital | 23 | 2.87 | 24 | 78 | 27 |
| Right Superior Frontal Gyrus | 22 | 3.08 | 42 | 78 | 67 |
| Left Planum Polare/insula | 21 | 2.97 | 77 | 47 | 43 |
| Posterior Cingulate | 20 | 2.94 | 48 | 47 | 57 |

| | | | | | |
|---|---|---|---|---|---|
| Posterior Cingulate | 20 | 2.85 | 43 | 43 | 58 |

**Post –hoc tests with trait anxiety**

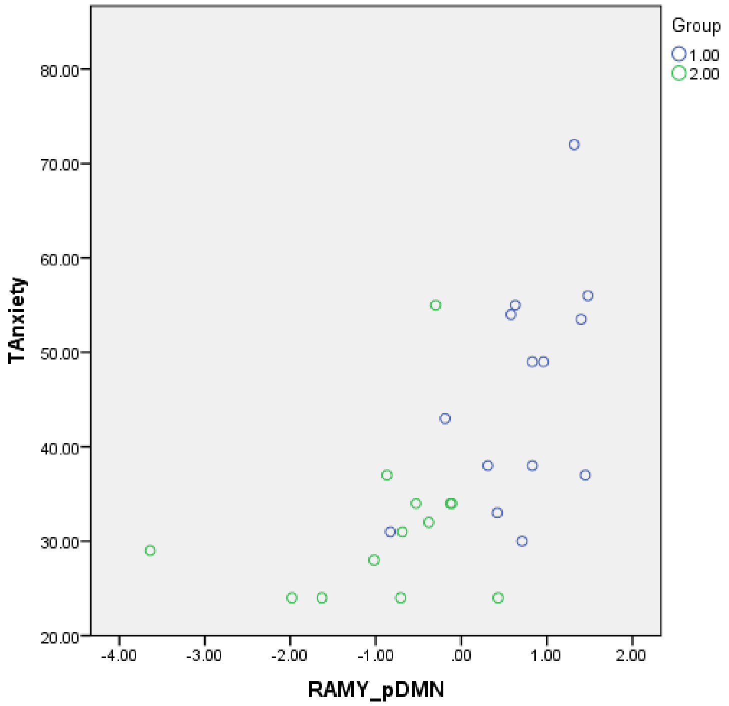


Group 1= opioid using participants, Group 2= control participants

**Figure 5a: Relationship between Trait anxiety and right amygdala-pDMN connectivity**

This figure shows extracted z scores for the connection pair plotted against trait anxiety scores.

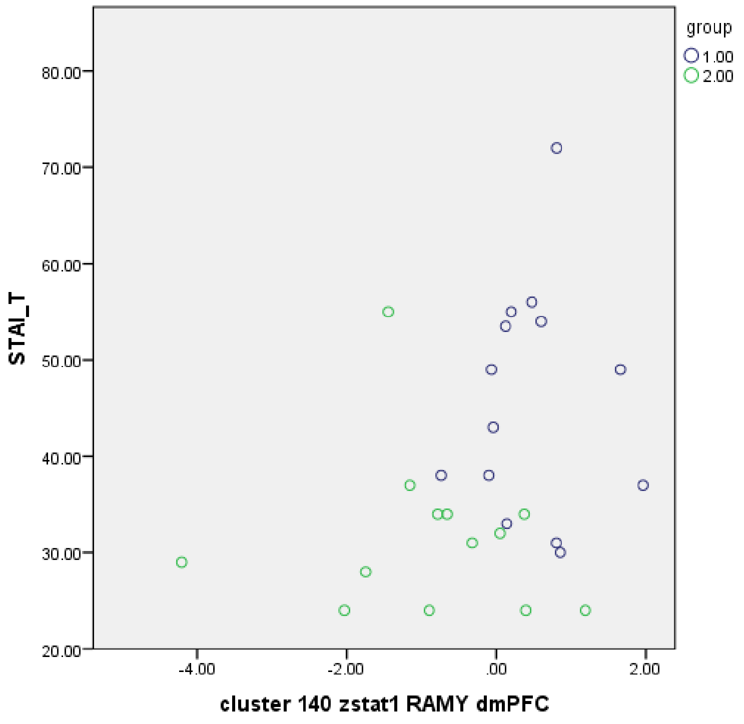


Group 1= opioid using participants, Group 2= control participants

**Figure 5b: Relationship between Trait anxiety and right amygdala-dmPFC connectivity**

This figure shows extracted z scores for the connection pair plotted against trait anxiety scores.

**Control participants > opioid using participants**

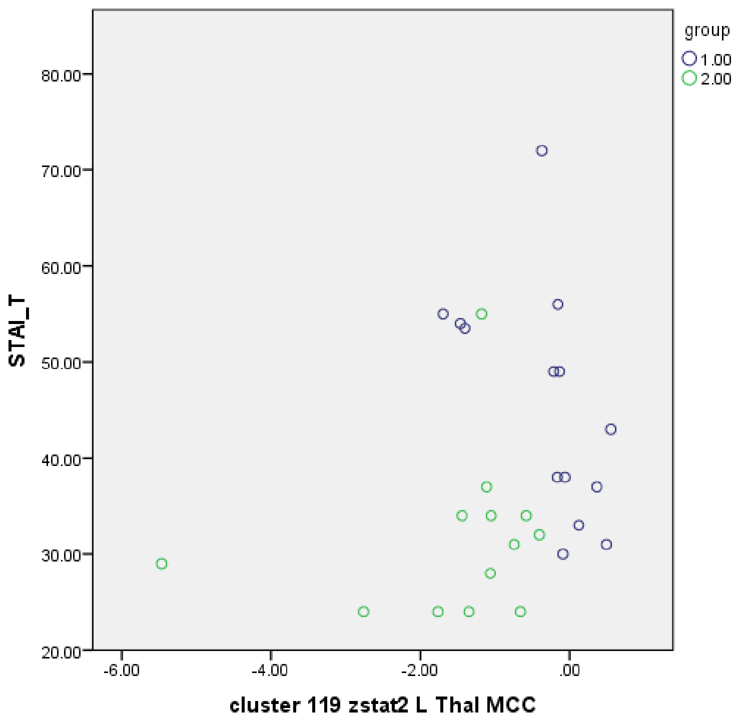


Group 1= opioid using participants, Group 2= control participants

**Figure 5c: Relationship between Trait anxiety and left thalamic-mid cingulate connectivity**

This figure shows extracted z scores for the connection pair plotted against trait anxiety scores.

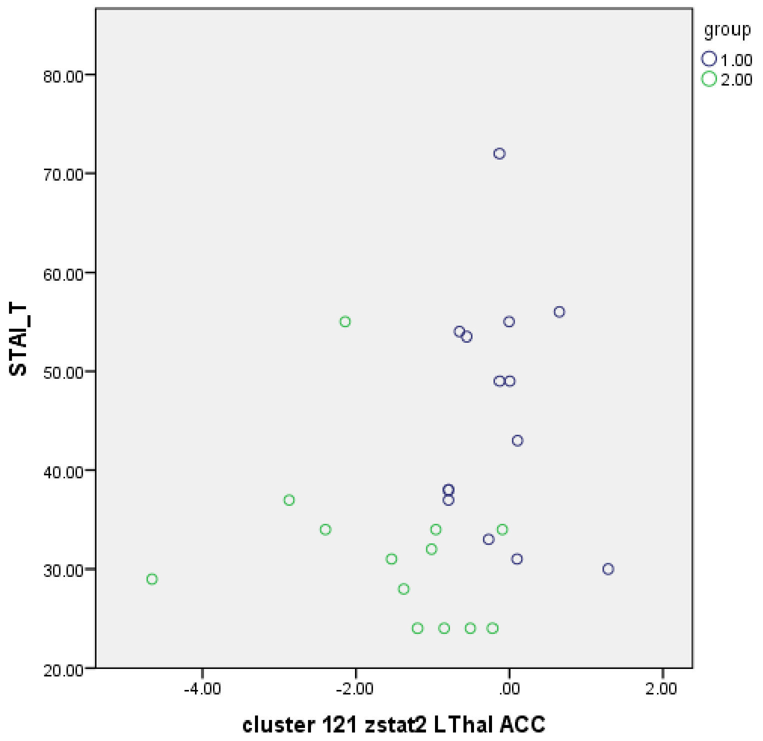


Group 1= opioid using participants, Group 2= control participants

**Figure 5d: Relationship between Trait anxiety and left thalamic-anterior cingulate connectivity**

This figure shows extracted z scores for the connection pair plotted against trait anxiety scores.
